# Association between methylenetetrahydrofolate reductase gene C677T polymorphism and susceptibility to polycystic ovary syndrome

**DOI:** 10.1101/2020.06.15.20132324

**Authors:** Vandana Rai, Pradeep Kumar

**Affiliations:** Department of Biotechnology, VBS Purvanchal University, Jaunpur -222003, India

**Keywords:** Polycystic Ovary Syndrome, PCOS, PCOD, MTHFR, C677T, meta-analysis

## Abstract

Polycystic ovary syndrome (PCOS) is the most common form of endocrinopathy of women. Several studies have investigated the association of methylenetetrahydrofolate reductase (MTHFR) gene C677T polymorphism with PCOS risk but the results are contradictory. So, the aim of the present study was to carry out a meta-analysis of a published case control studies to find out exact association between MTHFR gene C677T polymorphism and PCOS susceptibility. Pubmed, Springer link, Science Direct and Google Scholar databases were searched for case-control studies. Odds ratios (ORs) with 95% confidence intervals (CIs) was used as association measure and meta-analysis was performed using MIX and MetaAnalyst programs.

Meta-analysis of 24 studies showed strong significant association between C677T polymorphism and PCOS risk (for T vs. C: OR= 1.18, 95% CI=1.01-1.38, p=0.03; for TT vs. CC: OR= 1.37, 95% CI=1.0-1.89, p= 0.045; for TT + CT vs. CC: OR= 1.31, 95% CI= 1.07-1.62, p= 0.008; for CT vs. CC: OR= 1.31, 95% CI= 1.04-1.62, p= 0.01 and for TT vs. CT + CC: OR= 1.10, 95% CI= 0.82-1.47, p= 0.04). In subgroup analysis, MTHFR C677T polymorphism is significantly associated with PCOS risk with Asian individuallas but in Caucasian population MTHFR C677T polymorphism was not significantly associated with PCOS risk. In conclusion, C677T polymorphism is a risk factor for PCOS.

## Introduction

Polycystic ovary syndrome (PCOS) also known as polycystic ovary disease (PCOD) is the most common and widespread form of endocrinopathy of women of fertile, affecting up to 2 - 25% of women in reproductive age (Wijeyaratne et al., 2002). PCOS is characterized by oligomenorrhea or amenorrhea and hyperandrogenism (Fauser et al., 2004). It is associated with increased risk of pregnancy loss (Chang 2014). PCOS is a mutimetabolic syndrome and a risk factor for the obesity, type 2 diabetes, hypertension and dyslipidaemia (Wild et al., 1985; Talbott et al., 1995; Solomon, 1999). According to Rotterdam consensus (2003) diagnostic criteria, minimum two symptoms of the following should be present 1) clinical hyperandrogenism 2) oligoanovulation and 3) polycystic ovaries (PCO).

PCOS is a heterogeneous condition and earlier evidences and reports have suggested that the disturbed homocysteine (Hcy) methionine cycle increases concentration of homocysteine in the women, and higher concentration of homocystein is a risk factor for PCOS. Environmental and genetic factors play an important role in Hcy synthesis. Hcy is a sulfur containing amino acid and is converted into methionine in folate and methionin cycle. Methylenetetrahydrofolate reductase (MTHFR) enzyme is involved in the folate dependent remethylation of Hcy to methionine (Wu et al. 2012) and single nucleotide polymorphism (SNP) in MTHFR gene is the most common genetic defect of homocysteine metaboism. It catalyzes the conversion of 5,10-methylenotetrahydrofolate into 5-methylenotetrahydrofolate, which provides a single carbon for the methylation of homocysteine to methionine (Wu et al. 2012). C677T polymorphism (rs1801133) involves cytosine-to-thymine substitution at the 677 position (C677T), leads to an alanine-to-valine substitution at position 222 in the MTHFR enzyme. C677T polymorphism leads to thermolability of MTHFR enzyme, resulting in decreased enzyme activity (Frosst et al.,1995; Jacques et al. 1996). Frequency of C677T polymorphism is greatly varies in different global regions (Schneider et al., 1998; Wilcken et al., 2003; Spiridonova et al., 2004; Rai et al., 2010, 2012; Yadav et al., 2018).

First of all, Glueck and colleagues (1999) have reported an association between MTHFR C677T polymorphism and PCOS. Since then, several reports are published regarding MTHFR C677T polymorphism as a risk factor for PCOS with positive (Wu et al., 2016; Jiao et al., 2018; Amuhtar and Amohaidi, 2019)as we as negative (Sills et al., 2001; Lee et al., 2003; Orio et al., 2003) results. Hence, the aim of the present study was to explore whether the MTHFR C677T polymorphism is associated with susceptibility to polycystic ovary syndrome (PCOS). In the present study, authors have performed a large-scale meta-analysis to find out the exact association between the MTHFR C677T polymorphism and PCOS by including data from studies that were published from 1999 until now

## Methods

### Article Search

Published articles examining the effect of the MTHFR gene C677T polymorphisms on the risk of PCOS were identified through electronic database searches in Pubmed, Science direct, Google scholar and Springer Link. Databases searches were done from January 1990 Electronic database searches were supplemented by manual searches of references of review and published research articles. The following terms were used: ‘‘methylenetetrahydrofoate reductase’’, ‘‘MTHFR,’’ ‘‘C677T’’, ‘‘polycystic ovary syndrome’’ ‘‘polycystic ovary disease’’, ‘‘PCOS’’, ‘‘PCOD’’, ‘‘polymorphism’’. Literature search included all languages. If any study incuded different sample groups, authors have considered each sample group as a separate study in the meta-analysis.

### Inclusion criteria

Studies included in our meta-analyses were based on the following criteria: (1) published in a peer-reviewed journal, (2) detailed description of the samples tested (including sample size, ancestry of samples); and (3) data published before December 31, 2019. Articles were excluded if: (1) sample was not independent, (2) incomplete data/information, (3) family-based studies and genome-wide association studies (4) review, letter to editors and book chapters etc.

### Data Extraction

Name of first author, ethnicity, country name, number of controls and cases, and number of MTHFR genotypes in PCOS cases and Controls were extracted from each selected article.

### Statistical Analyses

Meta-analysis was done according to the method of Rai et al (2014). Pooled ORs were estimated by using fixed effects (Mantel and Haenszel,1959) and random effects (DerSimonian and Laird,1986) model depending upon heterogeneity. When there is considerable heterogeneity between studies then the pooled OR is preferably estimated using the RE model (Zintzaras and Ioannidis, 2005). The between studies heterogeneity was calculated and was quantified using the I^2^ statistic (Higgins and Thompson,2002). Sensitivity analysis was performed to evaluate by removing the studies not in Hardy–Weinberg equilibrium (HWE). Control population of each study was tested for Hardy–Weinberg Equilibrium (HWE) using calculator available at http://ihg.gsf.de/cgi-bin/hw/hwa1.pl. Publication bias was assessed by Egger regression asymmetry test (Egger et a.,1997). P values were two-tailed with a significance level of 0.05. All statistical analyses were performed using computer programs Meta-Analyst (Wallace et al.,2013) and Mix version 1.7 (Bax et al.,2006).

## Results

### Literature Search

Electronic database search yielded 161 studies, after applying strict inclusion and exclusion criteria total 23 articles were found suitable for the inclusion in the present meta-analysis.. Figure 1 shows the different phases of the study selection. Information was collected from 22 articles that associated MTHFR gene C677T polymorphism and PCOS (Glueck et al., 1999; Sills et al., 2001; Tsanadis et al., 2002; Lee et al., 2003; Orio et al., 2003; Palep-Singh et al., 2007; Choi et al., 2009; Bayram et al., 2010; Karadeniz et al., 2010; Idali et al., 2012; Jain et al 2012; Kazeerooni et al., 2013; Jiang et al., 2015; Naghavi et al., 2015; Qi et al., 2015; Carlus et al., 2016; Geng et al., 2016; Ozegowska et al., 2016; Sazafarowska et al., 2016 ; Wu et al., 2016; Jiao et al., 2018; Amuhtar and Amohaidi, 2019) (Table 1). In two studies, authors (Palep-Singh et al., 2007; Carlus et al., 2016) included two group of samples.. So, in present meta-analysis, both dataset were included as separate study, and the number of included studies were twenty four.

**Table 1.** Details of 24 included studies.

| Study | Country | Control no. | Case no. | HWE |
| --- | --- | --- | --- | --- |
| Glueck et al., 1999 | USA | 234 | 41 | 0.22 |
| Sills et al., 2001 | USA | 18 | 36 | 0.35 |
| Tsanadis et al., 2002 | Greece | 35 | 30 | 0.98 |
| Lee et al.,2003 | Korea | 100 | 86 | 0.66 |
| Orio et al., 2003 | Italy | 70 | 70 | 0.15 |
| Palep-Singh et al., 2007 | UK | 9 | 21 | 0.36 |
| Palep-Singh et al., 2007 | UK | 16 | 25 | 0.61 |
| Choi et al., 2009 | Korea | 115 | 227 | 0.06 |
| Bayram et al.,2010 | Turkey | 28 | 30 | 0 |
| Karadeniz et al., 2010 | Turkey | 70 | 86 | 0 |
| Idali et al., 2012 | Iran | 100 | 71 | 0.42 |
| Jain et al.,2012 | India | 95 | 92 | 0.36 |
| Kazerooni et al., 2013 | Iran | 60 | 120 | 0.007 |
| Jiang et al.,2015 | China | 122 | 90 | 0.36 |
| Naghavi et al.,2015 | Iran | 196 | 112 | 0.14 |
| Qi et al.,2015 | China | 58 | 115 | 0.14 |
| Carlus et al2016,Indo European | India | 100 | 93 | 0.36 |
| Carlus et al.,2016,Dravidian | India | 156 | 168 | 0.58 |
| Geng et al.,2016 | China | 236 | 175 | 0.06 |
| Ozegowska et a.,2016 | Poland | 99 | 168 | 0.49 |
| Sazafarowska et al., 2016 | Poland | 56 | 76 | 0.08 |
| Wu et al.,2016 | China | 257 | 244 | 0.23 |
| Jiao et al.,2018 | China | 307 | 336 | 0.11 |
| Almukhtar and Almohaidi,2019 | Iraq | 50 | 50 | 0.77 |

**Figure 1.**
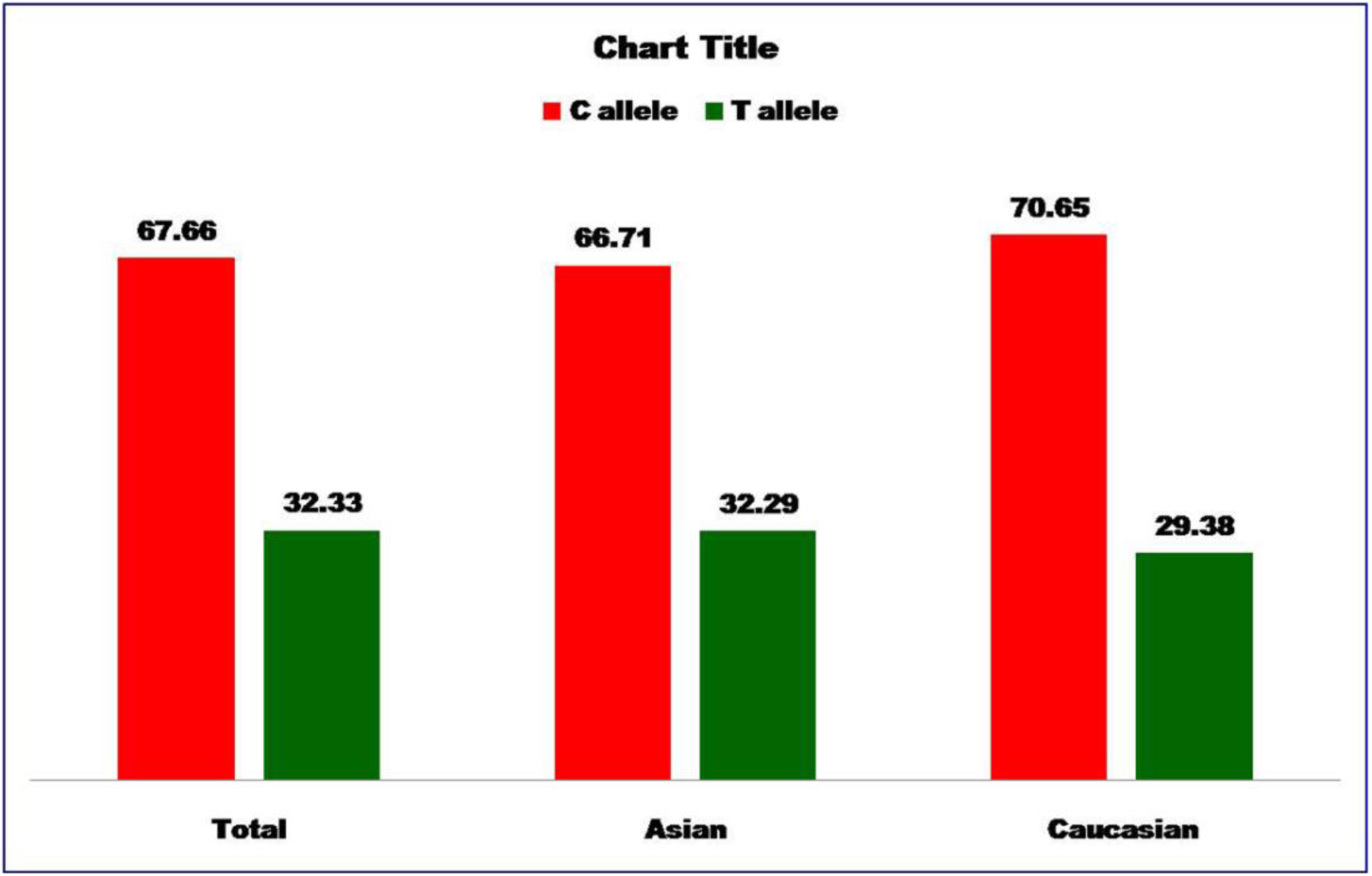
**Bar diagram showing percentage of C and T allele frequencies in control group of total 24 studies, 16 Asian studies and 8 Caucasian studies**.

### Characteristic of Eligible Studies

The included studies were published between 1999 and 2019. A total of 2562 cases and 2587controls were analyzed. The analysis included Caucasian samples in 10 studies and Asian samples in 14 studies (Table 1).

The prevalence of homozygotes (CC) among PCOS cases and healthy controls were 41.64% and 49.67%, respectively. The prevalence of heterozygotes (CT) among PCOS patients and controls were 41.26% and 36.72%, respectively. The prevalence of mutant homozygote (TT) among PCOS cases and the controls were 17.45% and 14.15%, respectively. In the PCOS cases, C allele was the most frequent: 62.06% whereas the frequency of the T allele was 37.94%. In the control group, the prevalence of C allele was 67.67%, and the prevalence of T allele was 32.33% (Figure 2). In four studies (Bayram et al., 2010; Karadeniz et al., 2010; Kazeerooni et al., 2013; Ozegows**ka** et al., 2015), control genotypes were not in Hardy–Weinberg equilibrium (P<0.05). All genetic models; -allele contrast (T vs C) homozygote (TT vs CC), co-dominant (CT vs CC), dominant (TT+CT vs CC) and recessive (TT vs CT+CC) models were used to evaluate C677T polymorphism as PCOS risk.

**Figure 2.**
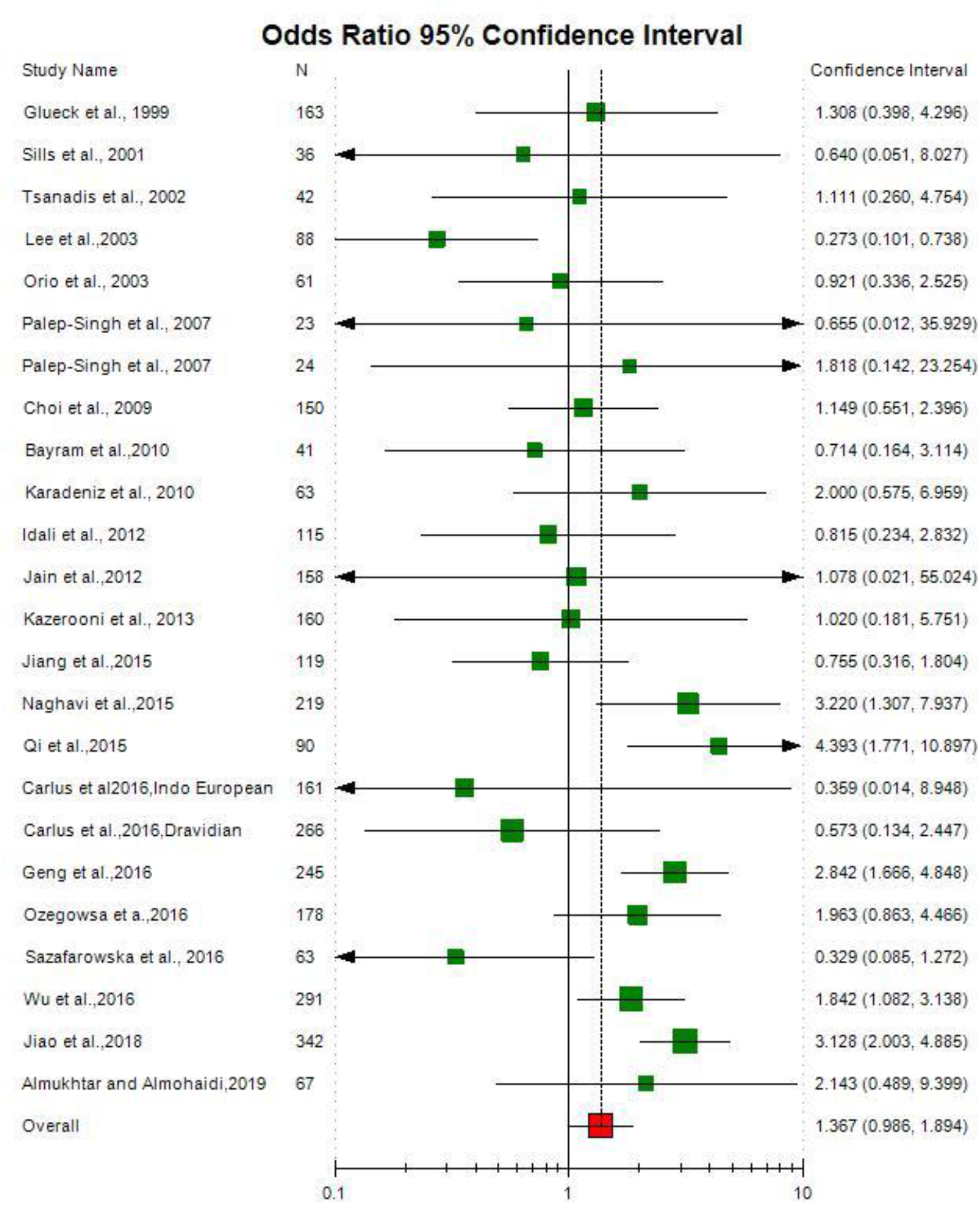
**Random effect Forest plot of homozygote model (TT vs. CC) of total 24 studies of MTHFR gene C677T polymorphism**.

### Meta-analysis

Table 2 summarizes the ORs with corresponding 95% CIs for association between T polymorphism and risk of PCOS in allele contrast, co-dominant, homozygote dominant, and recessive models. Meta-analysis of allele contrast (T vs. C) model showed significant association with both fixed effect (OR_TvsC_= 1.28; 95%CI= 1.17-1.40; p= <0.0001) and random effect model (OR_TvsC_= 1.18; 95%CI= 1.01-1.38; p= 0.03). Subjects with PCOS cases showed a significantly increased frequency of the T allele in comparison to control. (Table 2).

**Table 2:** **Summary estimates for the odds ratio (OR) of MTHFR C677T in various allele/genotype contrasts, the significance level (p value) of heterogeneity test (Q test), and the I^2^ metric and publication bias p-value (Egger Test)**.

| Genetic Models | Fixed effect<br>OR (95% CI), p | Random effect<br>OR (95% CI), p | Heterogeneity p-value (Q test) | I<sup>2</sup> (%) | Publication Bias (p of Egger's test) |
| --- | --- | --- | --- | --- | --- |
| Allele Contrast (T vs C) | 1.28(1.17-1.40),<0.0001 | 1.18(1.01-1.38),0.03 | <0.0001 | 61.1 | 0.07 |
| Co-dominant (Ct vs CC) | 1.34(1.17-1.59),<0.0001 | 1.31(1.04-1.62),0.017 | 0.0002 | 57.92 | 0.77 |
| Homozygote (TT vs CC) | 1.62(1.34-1.96),<0.0001 | 1.37(1.0-1.89),0.04 | 0.0019 | 53.06 | 0.05 |
| Dominant (TT+CT vs CC) | 1.39(1.23-1.58),<0.0001 | 1.31(1.07-1.62),0.008 | 0.0002 | 57.99 | 0.40 |
| Recessive (TT vs CT+CC) | 1.32(1.12-1.55),0.0007 | 1.10(0.82-1.47),0.04 | 0.001 | 55.06 | 0.01 |

Meta-analysis of twenty five studies showed an increased risk of PCOS among mutant homozygote variants (TT vs. CC; homozygote model), with both fixed (OR_TTvsCC_= 1.62; 95%CI= 1.34-1.96; p <0.0001) and random (OR_TTvsCC_= 1.39; 95%CI= 1.0-1.89; p= 0.04) effect models (Table 2, Figure 3). Association of heterozygous genotype (CT vs. CC; co-dominant model) was also observed significant with both fixed (OR_CTvsCC_= 1.34; 95%CI= 1.17-1.59; p <0.0001) and random (OR_CTvsCC_= 1.31; 95%CI= 1.04-1.62; p= 0.02) effect models (Table 2). Combined mutant genotypes (TT + CT vs.CC; dominant model) showed positive association with PCOS using both fixed (OR_TT+CTvsCC_= 1.39; 95%CI= 1.23-1.58; p <0.0001) and random (OR_TT+CTvsCC_= 1.31; 95%CI= 1.07-1.62; p= 0.008) effect models (Table 2). Similarly the recessive genotypes model (TT vs. CT + CC) also showed positive association with PCOS using both fixed (OR_TTvsCT+CC_= 1.32; 95%CI= 1.12-1.55; p= 0.0007) and random (OR_TTvsCT+CC_= 1.10; 95%CI= 0.82-1.47; p= 0.041) effect models (Table 2).

**Figure 3.**
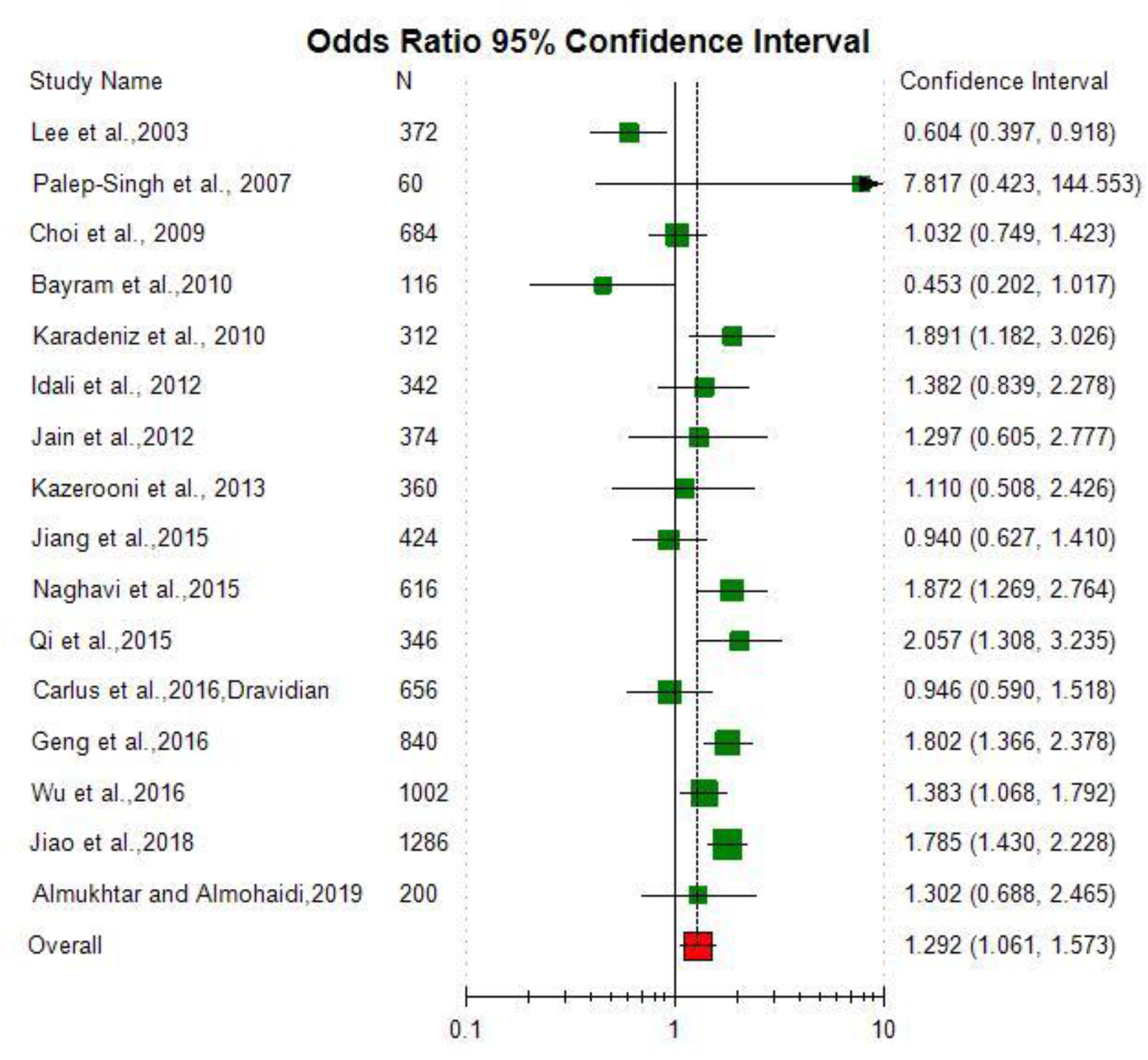
**Random effect Forest plot of allele contrast model (T vs. C) of total 16 Asian studies of MTHFR gene C677T polymorphism**.

### Sub-group Analysis

Out of 24 studies 16 studies (2023 cases and 1959 controls) were Asian region and 8 studies (539 cases and 628 controls) from Caucasian population. Meta-analysis of Asian studies revealed that except recessive model, a other four models showed significant association between MTHFR C677T polymorphism and PCCOS risk (for T vs. C: OR= 1.29, 95% CI=1.06-1.57, p=0.03; for TT vs. CC: OR= 1.38, 95% CI=0.91-2.11, p= 0.05; for TT + CT vs. CC: OR= 1.48, 95% CI= 1.13-1.92, p= 0.004; for CT vs. CC: OR= 1.49, 95% CI= 1,12-1.92, p= 0.005 and for TT vs. CT + CC: OR= 1.08, 95% CI= 0.74-1.58, p= 0.68) (Figure 4).

**Figure 4.**
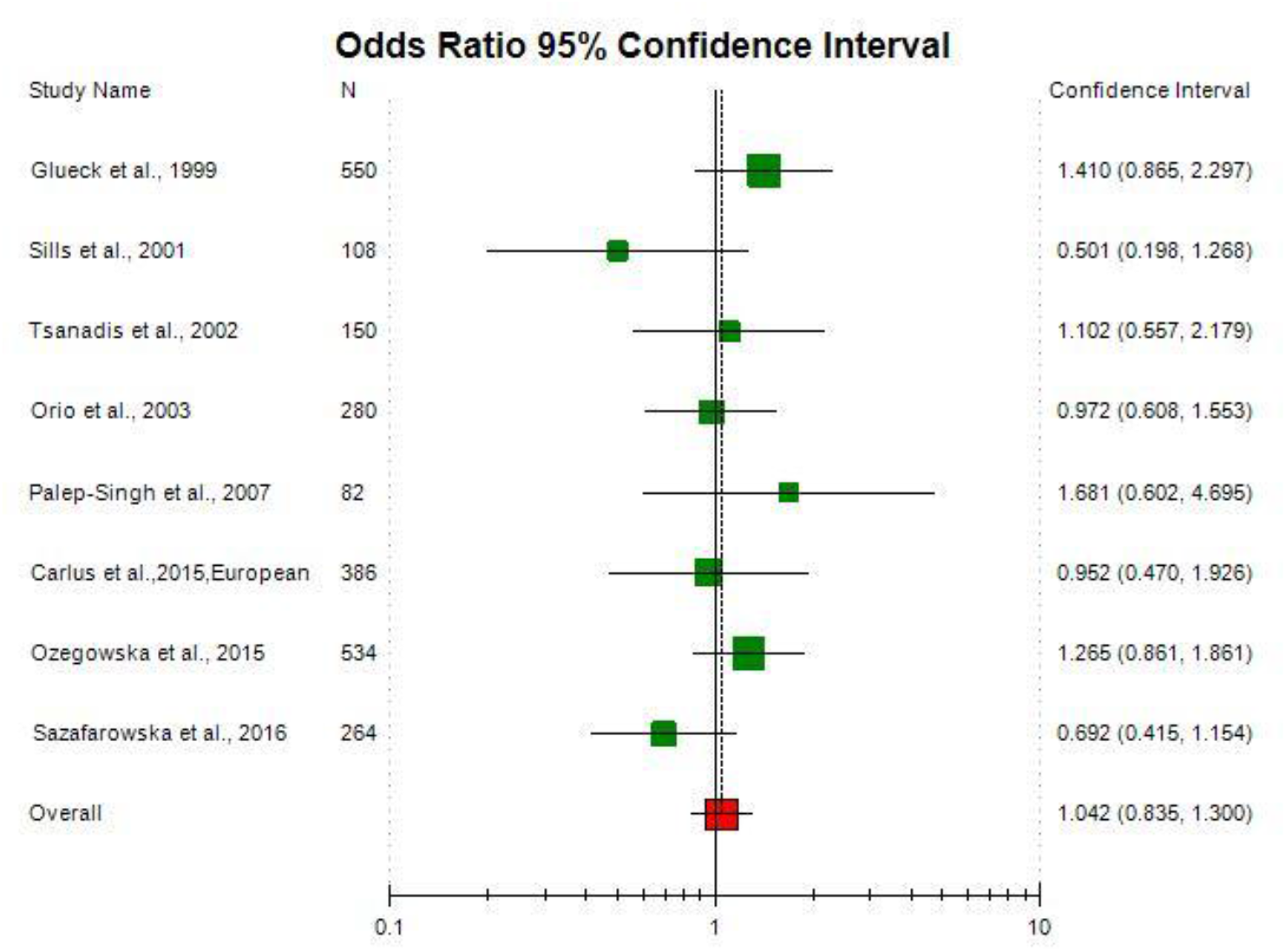
**Random effect Forest plot of allele contrast model (A vs. G) of total 8 Caucasian studies of MTHFR gene C677T polymorphism**.

Meta-analysis of Caucasian studies did not show any association between PCOS susceptibility and MTHFR C677T polymorphism (for T vs. C: OR = 1. 04, 95% CI= 0.83-1. 30, p= 0.31; for TT + CT vs. CC: OR = 1.07, 95% CI= 0.85-1.35, p= 0.54; for TT vs. CC: OR = 1.29, 95% CI= 0.87-1.91, p= 0.19) (Figure 5).

**Figure 5.**
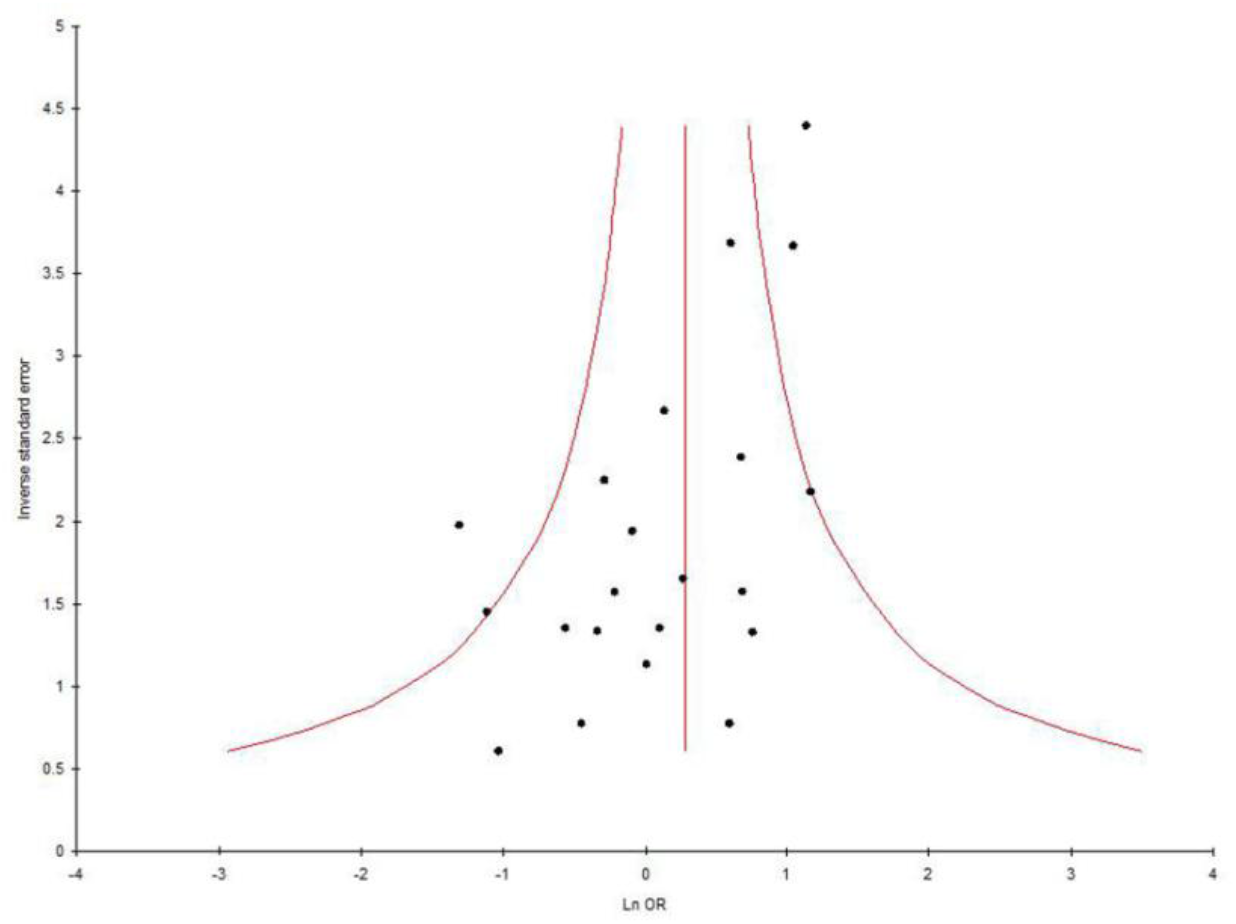
**Funnel plot-precision by log odds ratio for allele contrast model (T vs. C) of total 24 studies of MTHFR gene C677T polymorphism**.

### Heterogeneity analysis

A true heterogeneity existed between studies in allele contrast (P_heterogeneity_= <0.001, Q= 59.15, df= 24, I^2^= 61.12%, t^2^= 0.08), co-dominant (P_heterogeneity_= 0.0002, Q= 54.65, df= 24, I^2^= 57.92%, t^2^= 0.15), homozygote (P_heterogeneity_= 0.001, Q= 44.74, df= 24, I^2^= 53.06%, t^2^= 0.25) and dominant genotypes (P_heterogeneity_= 0.0002, Q= 54.74, df= 24, I^2^= 57.99%, t^2^= 0.14) comparisons. The ‘I^2^’ value was more than 50% in allele and a genotype comparisons, showed high level of true heterogeneity. Allele contrast meta-analysis of Asian studies showed significant higher heterogeneity (P_heterogeneity_= 0.0004, I^2^= 69.39%), but in Caucasian studies meta-anaysis true heterogeneity was absent (P_heterogeneity_= 0.34, I^2^= 11.06%).

### Sensitivity analysis

In sensitivity analysis, allele contrast meta-analysis was performed after exclusion of four studies (Bayram et al., 2010; Karadeniz et al., 2010; Kazeerooni et al., 2013; Ozegows**ka** et al., 2015) in which control population was not in HWE. Exclusion of four studies not in HWE did not affect heterogeneity and also did not increase the odds ratio.

### Publication bias

Precision and standard error Funnel plots are symmetrical and p-value of Eggers test, is higher than 0.05 (except recessive mode), showing the absence of publication bias (for T vs. C: P_Pb_= 0.07; for CT vs. CC: P_Pb_= 0.77; for TT vs. CC: P_Pb_= 0.05; for TT+CT vs. CC: P_Pb_= 0.41; and for TT vs. CT + CC:P_Pb_= 0.01) (Figure 6).

## Discussion

Several experimental and clinical evidences suggested that disturbances in folate (either deficiency or excess) and homocystein-methionine cycles resulted in ovarian dysfunctions-(i) ovulation in immature superovulated rats inhibited (Willmott et al., 1968; Wang et al., 2017), (ii) monkeys have degenerated Graffian follicles and an increase in atretic and cystic follicles (Mohanty and Das,1982; Wang et al., 2017) and (iii) homocysteine levels in PCOS cases and homocysteine levels returned to normal levels following folic acid supplementation (Bayraktar et al., 2004; Badawy et al., 2007; Kazerooni et al., 2007 ; Wang et al., 2017). In present meta-analysis, we tried to find out the exact associations between MTHFR C677T polymorphism and PCOS susceptibility. Our results indicated that the MTHFR polymorphism is risk factor for PCOS and OR is statistically significant (OR = 1.31; 95% CI= 1.07-1.62; p = 0.008).

Meta-analysis is a powerful strategy to find out the effects of genes in susceptibility of different diseases/disorders. In past two decades, numerous meta-analysis were published to evaluate the risk of genes polymorphism for different disease/ disorders-MTRR gene frequency (Yadav et al.,2019), down syndrome (Rai,2011; Rai et al., 2017; Rai and Kumar, 2018), cleft lip and palate (Rai, 2017), Gucose-6-phosphate dehydrogenas deficiency (Kumar et al.,2016), recurrent pregnancy loss (Rai,2014,2016), male infertility (Rai and Kumar, 2017), autism (Rai, 2016), schizophrenia (Rai et al., 2017), obsessive compulsive disorder (Kumar and Rai,2019), depression (Rai,2014,2017), epilepsy (Rai and Kumar,2018), Alzheimers disease (Rai, 2016), esophageal cancer (Kumar and Rai, 2018), prostate cancer (Yadav et al.,2016), breast cancer (Rai et al.,2017), endometrial cancer (Kumar et al., 2018) colorectal cancer (Rai, 2015), uterine leiomyioma (Kumar and Rai,2018),and ovary cancer (Rai, 2016).

Several limitations of this meta-analysis should be considered such as (i) crude OR calculated, (ii) presence of higher between heterogeneity, and (iii) included singe gene polymorphism. In addition to limitations, present meta-analysis also have several strengths like-absence of publication bias, inclusion of large number of studies as well as large sample size.

In conclusion, the results indicate that the MTHFR C677T polymorphism is risk factor for PCOS (OR = 1.31; p = 0.008). Subgroup analysis based on ethnicity also confirmed the results that MTHFR C677T polymorphism is risk factor for PCOS in Asians but not in Caucasian population. In future, Studies with larger sample sizes from different population of globe are required to come to conclusion regarding MTHFR C677T polymorphism and PCOS association.

## Data Availability

Yes all data available

## Compliance with Ethical Standards

### Conflict of Interest

None

Informed consent and Ethical Clearance are not required, as there was no human or animal involvent in the present study

### Sources of financial support

Nil

